# Antiviral use and the effects of drug resistance on the transmission dynamics of influenza

**DOI:** 10.1101/2025.03.10.25323668

**Authors:** Lea Schuh, Nikolaos I. Stilianakis

## Abstract

The effectiveness of antivirals in mitigating influenza outbreaks depends on both their ability to reduce the number of infections and the risk of drug resistance. We extended a previously developed mathematical model to investigate the impact of mitigation strategies, including mono or combination antiviral treatment or chemoprophylaxis and vaccination, on influenza transmission dynamics. Our findings indicate that chemoprophylaxis is more effective than treatment in reducing influenza burden, except when the resistant strain has a high transmission rate, in which case chemoprophylaxis may trigger a resistance-driven secondary infection wave. Combination therapy considerably reduces resistance emergence with similar infection numbers as mono-therapy. Vaccination coverage of at least 80% is required to prevent outbreaks; otherwise, antivirals can contribute to outbreak control provided drug resistance emergence is low. This analysis could inform public health decision-making by providing guidance on effective mitigation strategies for influenza outbreaks, considering their benefits against the risk of drug resistance.

## Introduction

Influenza poses a significant threat to global health, with approximately one billion cases of seasonal influenza and between 290,000 to 650,000 deaths annually, as reported by the World Health Organization (WHO) [1]. This highlights the urgent need for effective prevention and mitigation strategies against future influenza outbreaks, including vaccines, antivirals, and a range of public health measures. The WHO’s Global Influenza Strategy for 2019-2030 highlights the vital importance of antiviral drugs in the current preparedness and response efforts, particularly during the early stages of an outbreak, where timely access to therapeutics could be a critical mitigation factor [2]. Three primary classes of antiviral drugs are currently available: M2 inhibitors, such as amantadine and rimantadine, neuraminidase (NA) inhibitors, such as oseltamivir and zanamivir, and polymerase inhibitors, such as baloxavir marboxil. However, the general benefit of antivirals could be compromised by the emergence of resistant viral strains, rendering the effectiveness of antivirals limited. Notably, the widespread resistance of currently circulating influenza A strains to the M2 inhibitor amantadine has already led to a reduction of available therapeutic options [3].

The H5N1 avian influenza virus has recently raised new concerns, posing an increasing threat in terms of spillover and pandemic potential. As of 2024, the United States has reported 66 confirmed and 7 probable human cases of avian influenza, with the first recorded death in early January 2025 [4, 5]. Although human-to-human transmission has not been reported to date, the successful adaptability of the recent circulating H5N1 strain to mammalian transmission warrants continued surveillance and preparedness. In response to this potential emerging public health threat, the CDC offers influenza antivirals, particularly NA inhibitor oseltamivir, for both treatment and as post-exposure prophylaxis to prevent infection and continues with broader preparedness activities, including the planning for an H5 vaccination program, should one be needed [6, 7]. Similarly, the European Commission’s Health Emergency Preparedness and Response Authority (HERA) initiated a joint procurement framework contract for pre-pandemic vaccine doses demonstrating pro-active efforts to enhance preparedness against H5N1 [8].

Given the pivotal role of antiviral drugs in past influenza outbreaks, it is essential to optimize their use for emerging outbreaks. This necessitates a comprehensive approach to understanding the dynamics of influenza transmission and the impact and limitations of existing prevention and mitigation strategies, including the anticipation of emerging antiviral resistance.

Prior research has demonstrated the value of mathematical modeling in quantifying the dynamics of influenza transmission to better understand the impact of prevention and mitigation strategies, including vaccination and antiviral therapy. Research findings consistently showed that (post exposure) chemoprophylaxis yields a more effective use of antivirals, leading to a greater reduction in disease burden compared to a treatment-only approach [9, 10, 11, 12] and identified chemoprophylaxis to lead to similar outcomes than a population with a 80% vaccination coverage [13]. Mathematical modeling studies not only highlighted the potential benefits of widespread antiviral interventions but were also developed to explore different antiviral combination therapies, including sequential and early combination treatment [14] or chemoprophylaxis/treatment combinations [15], to evaluate the potential risks associated with the emergence of resistance. Alternatively to antivirals, vaccines offer a more longterm solution to influenza outbreaks. However, vaccine production against specific strains results in accessibility delays, particularly during the outbreak’s early stages. Mathematical models have also been used to optimize vaccination strategies during influenza outbreaks, examining key factors such as ideal vaccine dosage, timing, and target population demographics to inform effective immunization approaches [16, 17, 18, 19, 20].

Building on this, we developed a comprehensive mathematical model, including mono and combination treatment or chemoprophylaxis with antivirals oseltamivir or zanamivir and vaccination, to quantitatively describe the infection dynamics of an influenza outbreak under different prevention and mitigation strategies for (semi) closed systems, particularly relevant for vulnerable groups and settings, such as nursing homes and boarding schools. Our results provide insights into the potential benefits and limitations of antiviral therapy and could help inform public health decision-making to better prepare for and respond to future influenza outbreaks.

## Results

### Combination therapy reduces the emergence of drug resistance

We extended previous epidemiological models of influenza infections under antiviral mono treatment or chemo-prophylaxis to incorporate also combination treatment (Figures 1A and S1A) or chemoprophylaxis (Figures 1C and S1C) [9, 10].

**Figure 1.**
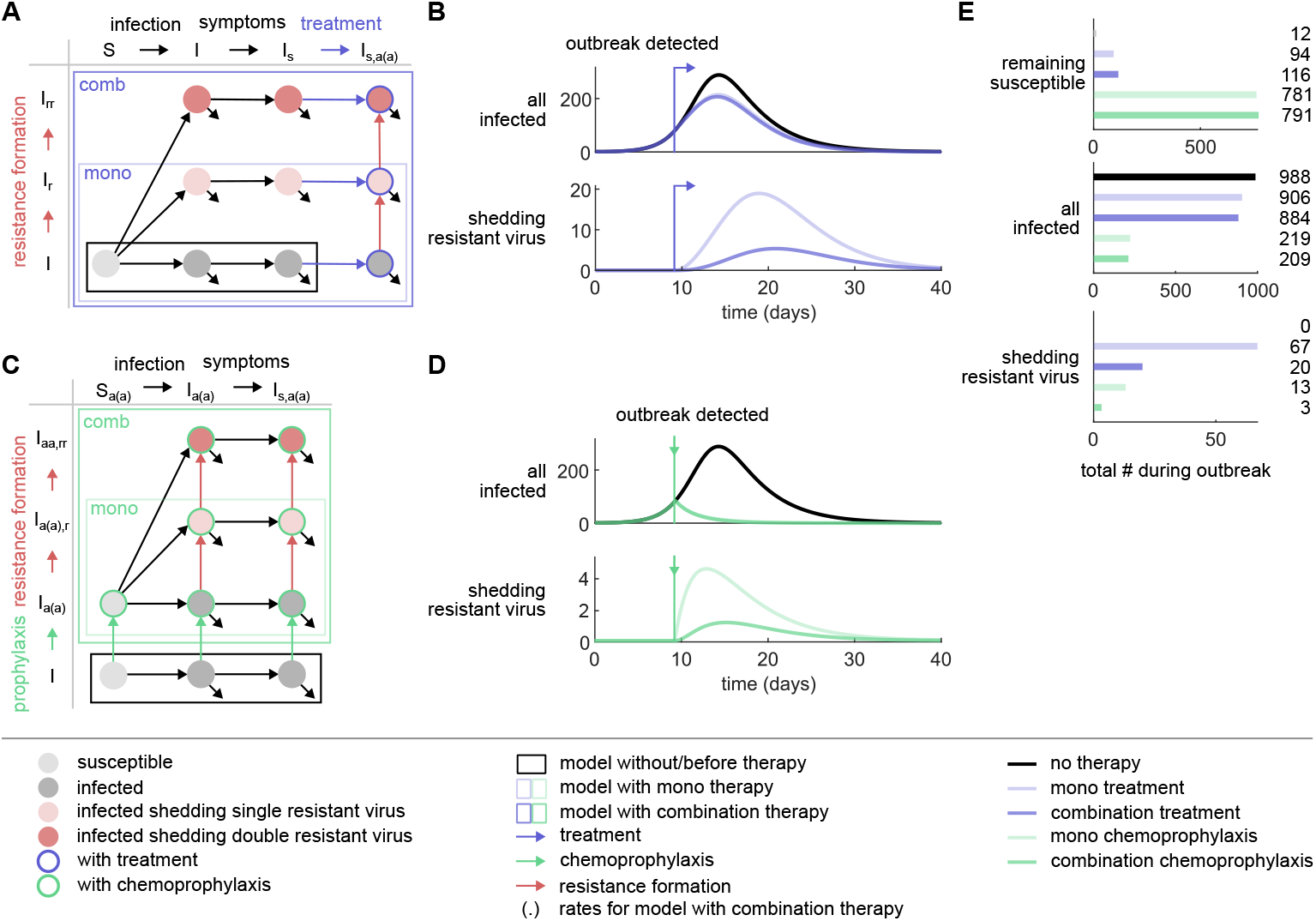
Influenza epidemiological model and simulated outbreak outcomes. (A) Schematic presentation of the influenza epidemiological model with antiviral mono and combination treatment. Susceptible (*S*) individuals become infected (*I*) and symptomatic (*I*_*s*_). Upon outbreak detection, symptomatic individuals are treated with antiviral mono or combination treatment (blue arrows, *I*_*s,a*(*a*)_), facilitating the development of single and/or double resistant virus (red arrows, *I*_*s,a*(*a*),*r*_ and *I*_*s,aa,rr*_). Infections with individuals shedding resistant virus lead to a transmission of the resistant virus (*I*_*r*_ and *I*_*r r*_). All infected individuals recover and become immune. (B) Numerical results of the model for infected individuals (top) and individuals shedding resistant virus (bottom) under different treatment scenarios: no treatment (black), mono treatment (light blue), and combination treatment (dark blue). The arrow indicates the time of outbreak detection and, consequently, treatment initiation. (C) Schematic presentation of the influenza epidemiological model with antiviral mono and combination chemoprophylaxis. All individuals are treated with antiviral mono or combination chemoprophylaxis (green arrows, *S*_*a*(*a*)_, *I*_*a*(*a*)_, and *I*_*s,a*(*a*)_) upon outbreak detection. This facilitates the formation of single or double resistant virus (red arrows, *I*_*a*(*a*),*r*_, *I*_*s,a*(*a*),*r*_ and *I*_*aa,rr*_, *I*_*s,aa,rr*_). All infected individuals recover and become immune. (D) Simulation of the influenza epidemiological model for infected individuals (top) and individuals shedding resistant virus (bottom) under different chemoprophylaxis scenarios: no chemoprophylaxis (black), mono chemoprophylaxis (light green), and combination chemoprophylaxis (dark green). The arrow indicates the time of outbreak detection and chemoprophylaxis administration. (D) Outcome of the outbreak for the remaining susceptible individuals (top), infected individuals (middle), and individuals shedding resistant virus (bottom) under different therapy scenarios: no therapy, mono treatment, combination treatment, mono chemoprophylaxis, and combination chemoprophylaxis. Same color-coding as in (B) and (D).

The new models allowed us to simulate the spread of influenza under various therapy scenarios (Figures 1B and D and Figure S1B and D) and compare their outcomes (Figures 1E and S1E). Numerical results show that the initiation of treatment upon outbreak detection resulted in a slower increase of infected individuals compared to the scenario without treatment. However, it did not completely prevent new infections (Figure 1B). In contrast, the introduction of antiviral chemoprophylaxis led to an immediate reduction in the number of infected individuals (Figure 1D). Consistent with previous findings, our numerical results showed that the remaining number of susceptible individuals after the outbreak was highest in scenarios with prophylactic measures, with a prevention rate of 78% for mono chemoprophylaxis (Figure 1E). In contrast, the prevention rate for mono treatment was considerably lower, at 8%. Moreover, the number of individuals shedding resistant virus is considerably lower in the scenario with mono chemoprophylaxis than for mono treatment, while the percentages of individuals shedding resistant virus are comparable across both therapy scenarios (6% and 7%, respectively). While combination therapy did not lead to a notable reduction in the numbers of infections compared to mono therapy for both treatment and chemoprophylaxis it did considerably reduce the absolute numbers and percentages of individuals shedding resistant virus (from 67 to 20, that is an enhanced reduction by 70%, for treatment, and from 13 to 3, that is an enhanced reduction by 77%, for chemoprophylaxis) and percentages (from 7% to 2% for treatment and from 6% to 1% for chemoprophylaxis). Overall, the implementation of chemoprophylaxis control measures yielded the most favorable outcomes in terms of reducing the spread of infection. The use of antiviral combination therapy contributed mainly to reduce the emergence of resistant viral strains.

### Resistance is limited even for high de novo formation

Each season influenza strains display distinct characteristics that may change their potentials to form resistance. Using the previously introduced models, we investigated the effects of varying de novo resistance formation rates on the outcomes of influenza outbreaks under different therapy scenarios. In line with results observed under standard model parameters (Table 2), we found that the number of infections remains lower with chemoprophylaxis relative to treatment, and lower still with combination therapy as opposed to mono therapy, regardless of changes in relative de novo resistance formation rates (Figures 2A and S2A-D, top).

**Figure 2.**
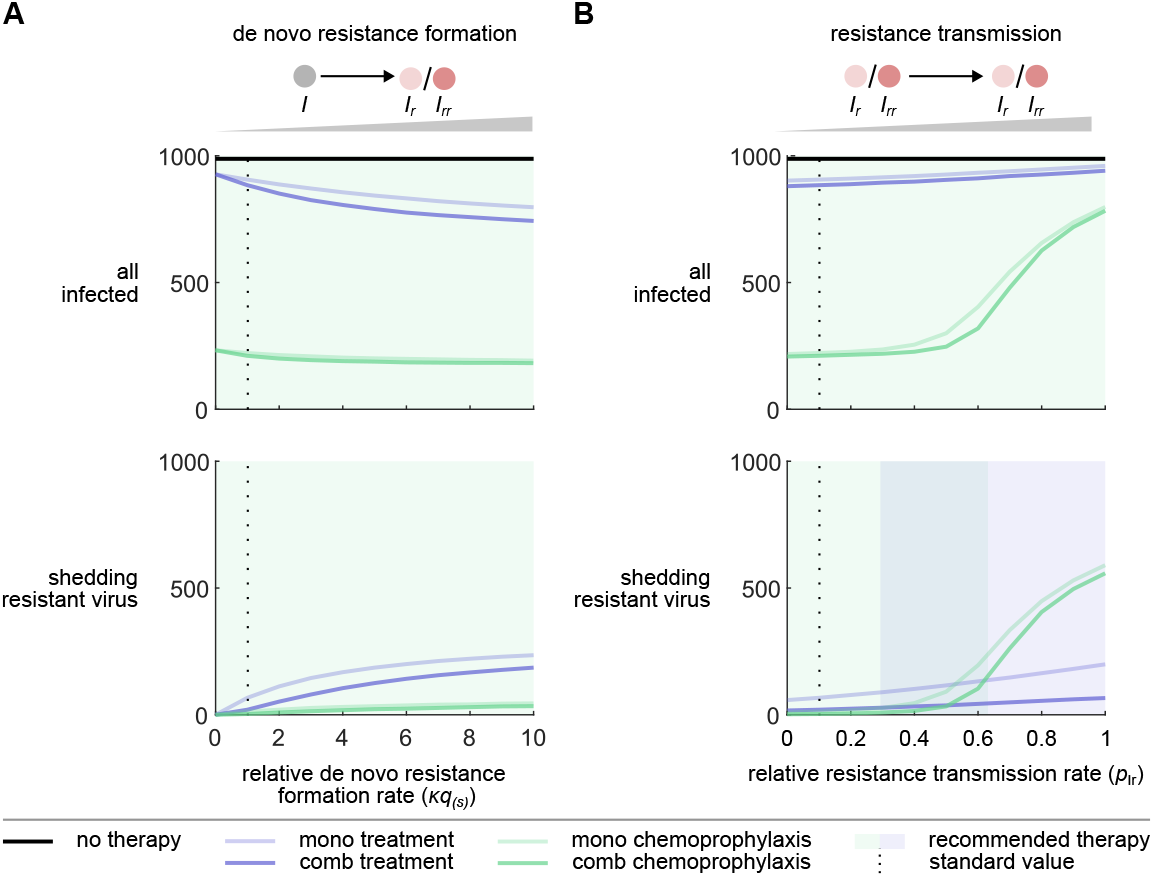
The optimal therapy depends on the resistance formation rates and optimization objective. (A) Increasing the relative de novo resistance formation rate leads to higher but converging numbers of infected individuals (top) and those shedding resistant virus (bottom). This trend is observed for the therapy scenarios of mono treatment (light blue), combination treatment (dark blue), mono chemoprophylaxis (light green), and combination chemoprophylaxis (dark green). The number of infected individuals without therapy is shown by a solid black line. The standard value of the de novo resistance formation rate is set to 1 and indicated by a dotted black line. The optimal therapy with respect is highlighted by the background color. (B) Increasing the relative resistance transmission rate of the resistant virus with respect to the wildtype virus leads to increasing numbers of infected individuals (top) and those shedding resistant virus (bottom). While linear increases in the numbers are observed for mono and combination treatment, mono and combination chemoprophylaxis result in sigmoidal increases. The number of infected individuals without therapy is shown by a solid black line. The standard value of the relative resistance transmission rate is set to 0.1 and indicated by a dotted black line. Same color-coding as in (A).

Interestingly, an increase in the relative de novo resistance formation rate leads to a decrease in the total number of infections. The underlying reason for this phenomenon is that the development of resistance involves the accumulation of mutations, which impairs the virus’s fitness. As a result, when de novo resistance increases, a larger proportion of individuals sheds virus with reduced transmissibility, ultimately contributing to a decrease in the overall number of infected individuals. Meanwhile, a rise in the de novo resistance formation rate leads to a greater number of individuals shedding resistant virus for all examined therapy scenarios, with consistently lower numbers for chemoprophylaxis (Figures 2A and S2A-D, bottom). The change in the number of individuals shedding resistant virus becomes less for larger de novo resistance rates, suggesting that other factors than de novo resistance formation might be more critical for the successful spread of resistance within a population. Accordingly, we did not observe simulations with a secondary infection wave driven by the resistant strain for any of the therapy scenarios. To quantify this result, we derived and computed the control reproduction numbers, which here quantifies the average number of individuals shedding resistant virus that would arise from introducing a single infected individual into an entirely susceptible population (Methods section). Our calculations demonstrate that, under the current model parameters, the effect of an increasing relative de novo resistance formation rate on the number of individuals shedding resistant virus is limited and will never result in a secondary infection wave driven by the resistant strain (Figure S2E). Here, we only considered the effects of de novo resistance formation on the spread of influenza at the population level. However, a high de novo resistance formation rate can strongly impact the individual’s disease progression. Overall, chemoprophylaxis outperforms treatment across varying relative de novo resistance formation rates.

### High transmission rates of the resistant strain can lead to secondary infection waves

Similarly, seasonal influenza strains can differ in their capacity to transmit resistance. In our standard model, we assumed that acquiring mutations enabling resistance comes at a fitness cost to the virus, resulting in a reduced transmission rate for the resistant strain. Using the previously established models, we investigated the effects of varying resistance transmission rates on the outcomes of influenza outbreaks under different therapy scenarios. We found that increasing the resistance transmission rate of the resistant viral strain relative to the wildtype virus resulted in a rise in the total number of infections, with consistently lower infections under chemoprophylaxis scenarios compared to treatment and even lower numbers for combination compared to mono therapy (Figures 2B and S2F-I, top). The patterns of increase in infections, however, were notably different between treatment and chemoprophylaxis. Treatment resulted in only a modest and linear growth in infection numbers as the relative resistance transmission rates increased. In contrast, for chemoprophylaxis, once the relative transmission rates exceeded 0.5, a secondary wave of resistance-driven infections emerged, causing a sharp and sigmoidal increase in the number of infections. This pattern was also reflected in the number of individuals shedding the resistant virus (Figures 2B and S2F-I, bottom). The control reproduction numbers confirm this numerically, where relative resistance transmission rates above 0.5 lead to increased infection numbers for chemoprophylaxis (Figure S2J). At the same time, for treatment, the control reproduction numbers demonstrate that not sufficient susceptible individuals remain upon treatment initiation allowing for a resistance-driven secondary infection wave (Figures S2J and S2K). These observations suggest that while resistance transmission modestly affects infection dynamics during treatment, it has a considerable impact during chemoprophylaxis. Nevertheless, chemoprophylaxis is still the preferred therapy when aiming to reduce the overall number of infections. However, when the goal is to limit the development of resistance, chemoprophylaxis is only advantageous for lower relative resistance transmission rates. At higher rates, treatment becomes the better option. Overall, our findings suggest that the bottleneck for the emergence of a secondary infection wave driven by the resistant strain is the reduced fitness of the resistant virus, not the formation of resistance.

### Vaccination in combination with treatment is more effective than with chemoprophylaxis

We extended the epidemiological models for influenza infections with mono treatment or mono chemoprophylaxis to also include vaccination (Figure 3A and C and Figure S3A and C) [9, 10].

**Figure 3.**
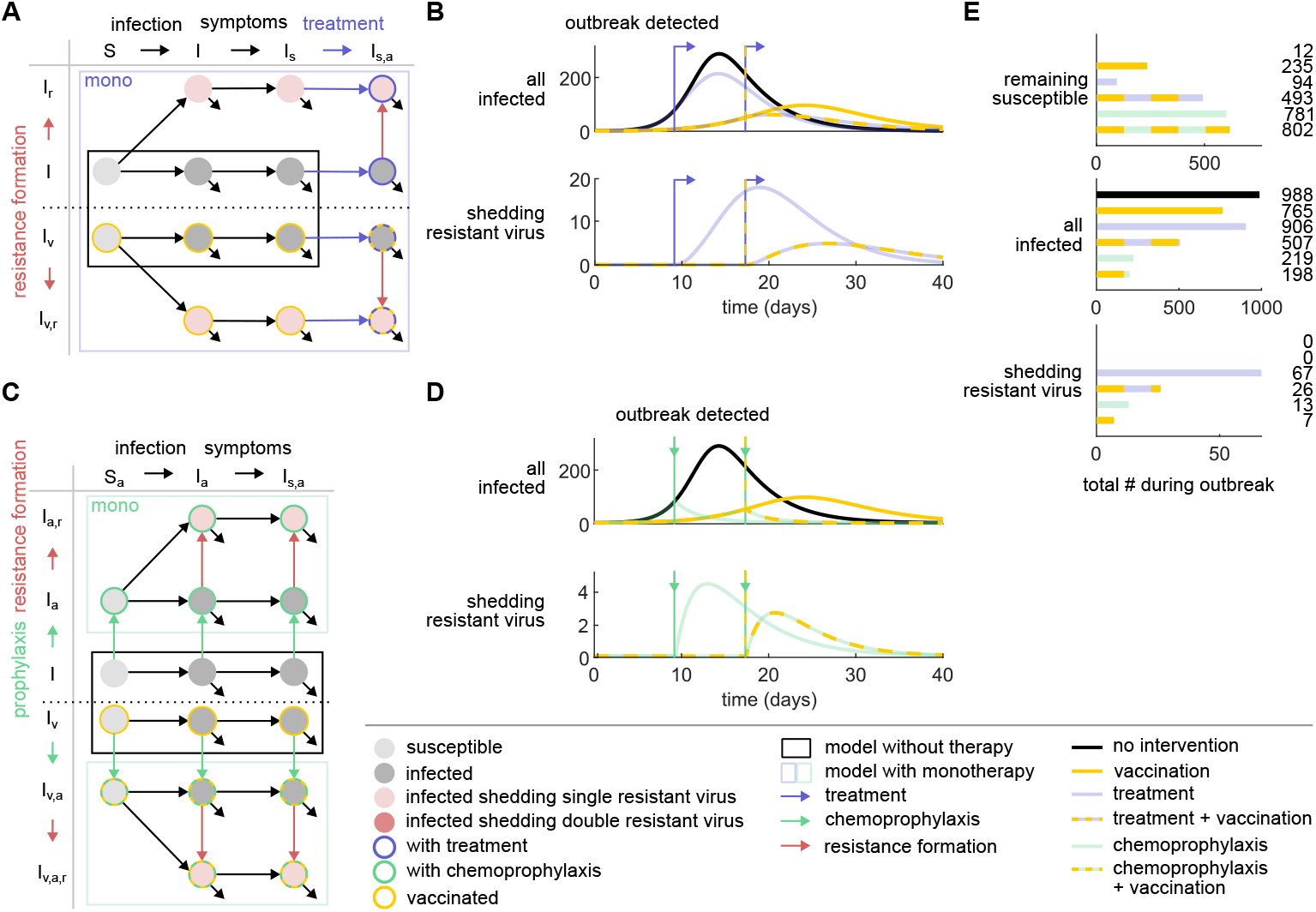
Vaccination is most effective in combination with treatment to reduce infections and resistance. (A) Schematic presentation of the influenza epidemiological model with a vaccinated subpopulation and treatment + vaccination. Non-vaccinated and vaccinated susceptible individuals (*S* and *S*_*v*_, respectively) become infected (*I* and *I*_*v*_, respectively) and symptomatic (*I*_*s*_ and *I*_*v,s*_, respectively). Upon outbreak detection, all symptomatic individuals are treated with antiviral treatment (blue arrows, *I*_*s,a*_ and *I*_*v,s,a*_, respectively), facilitating the development of resistant virus (red arrows, *I*_*s,a,r*_ and *I*_*v,s,a,r*_, respectively). Infections with individuals shedding resistant virus lead to a transmission of the resistant virus (*I*_*r*_ and *I*_*v,r*_). All infected individuals recover and become immune. (B) Simulations of the influenza epidemiological model with different intervention scenarios: no intervention (black), vaccination (yellow), treatment (blue), and treatment + vaccination (blue-yellow). The blue arrow indicates the times of outbreak detection and, consequently, treatment initiations. (C) Schematic presentation of the influenza epidemiological model with antiviral mono chemoprophylaxis. All individuals are treated with antiviral chemoprophylaxis (green arrows, *S*_*a*_, *I*_*a*_, and *I*_*s,a*_ and *S*_*v,a*_, *I*_*v,a*_, and *I*_*v,s,a*_, respectively) upon outbreak detection. This facilitates the formation of resistant virus (red arrows, *I*_*a,r*_, *I*_*s,a,r*_ and *I*_*v,a,r*_, *I*_*v,s,a,r*_, respectively). (D) Simulation of the influenza epidemiological model with different intervention scenarios: no intervention (black), vaccination (yellow), chemoprophylaxis (green), and chemoprophylaxis + vaccination (green-yellow). The green arrow indicates the times of outbreak detection and chemoprophylaxis administrations. (E) Outcome of the outbreak under different intervention scenarios: no intervention, vaccination, treatment, treatment + vaccination, chemoprophylaxis, and chemoprophylaxis + vaccination. Same color-coding as in (B) and (D).

Using the extended models, we simulated the spread of influenza under various intervention strategies, including vaccination, treatment + vaccination (Figures 3B and S3B), and chemoprophylaxis + vaccination (Figures 3D and S3D). While vaccination serves to flatten the curves for the number of infections and of individuals shedding resistant virus, it also extends the duration of the outbreak. Vaccination increases the number of remaining susceptible individuals (Figures 3E and S3E). The percentage of individuals spared from infection due to vaccination is higher for the intervention of treatment (44%) and lower for the intervention of chemoprophylaxis (10%), while chemoprophylaxis + vaccination results in the lowest number of individuals shedding resistant virus. Overall, our results demonstrate that the joint effects of vaccination and treatment are more effective than of vaccination and chemoprophylaxis.

### A vaccination coverage of 80% prevents outbreaks

Before each influenza season, individuals especially those who belong to risk groups such as elderly are called upon to get vaccinated. Despite these efforts and a target vaccination coverage of 75%, just over half (51%) of the people aged 65 years and older were vaccinated against influenza in the European Union in 2021. Using the previously introduced models, we investigated how varying vaccination coverages and their joint intervention with antiviral mono therapies may lead to different outcomes (Figures 4A and S4A-C).

**Figure 4.**
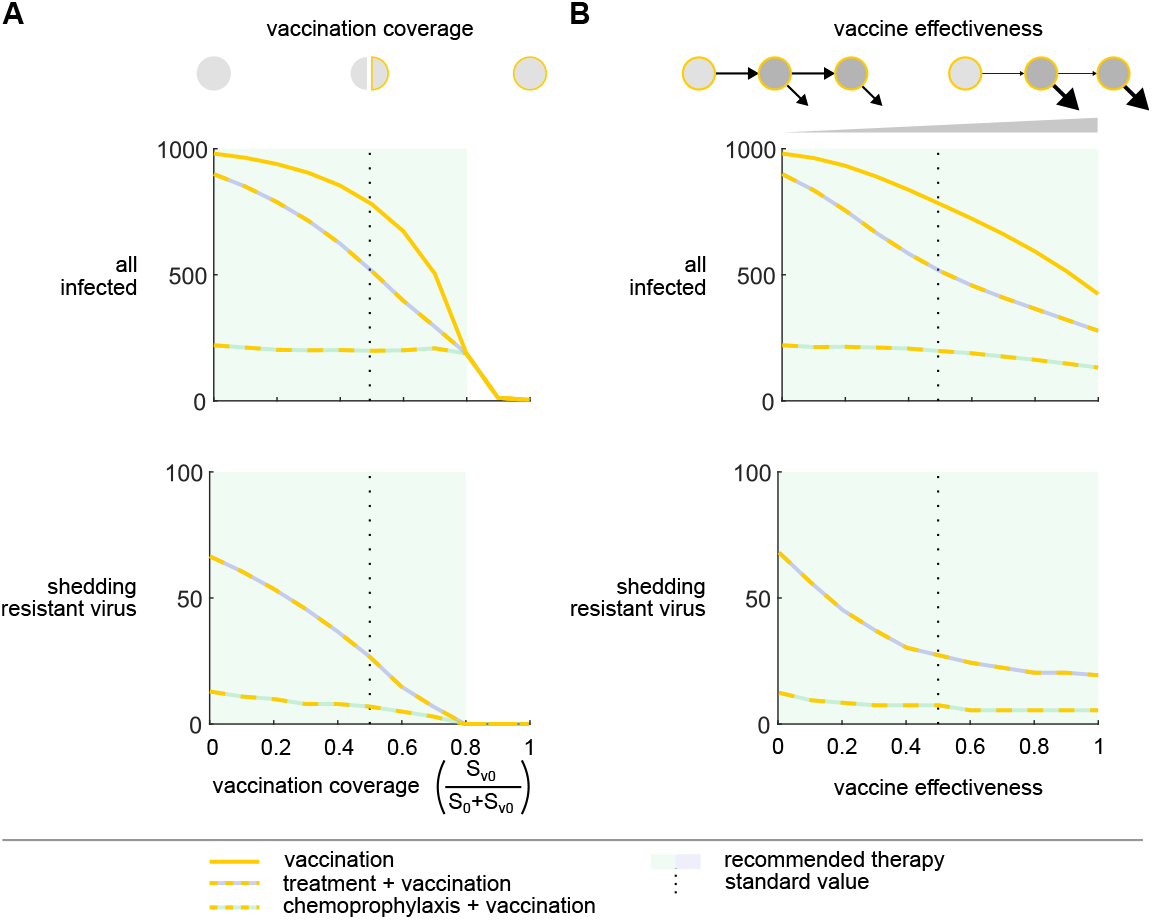
80% or more of vaccinated individuals prevents the influenza outbreak. (A) Increasing the vaccination coverage leads to lower numbers of infected individuals (top) and those shedding resistant virus (bottom) for the intervention scenarios of vaccination (yellow) and treatment and vaccination (blue-yellow). For chemoprophylaxis and vaccination (greenyellow) only the number of individuals shedding resistant virus decreases with an increasing vaccination coverage. The standard value of the vaccination coverage is 0.5 and indicated by a dotted black line. The optimal therapy is highlighted by the background color. (B) Increasing the effectiveness of the vaccine leads to lower numbers of infected individuals (top) and those shedding resistant virus (bottom) for the intervention scenarios of vaccination, treatment and vaccination, and chemoprophylaxis and vaccination. The standard value of the effectiveness of the vaccine is set to 0.5 and indicated by a dotted black line. Same color-coding as in (A).

Increasing vaccination coverages 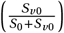 lead to less overall infections for vaccination and treatment + vaccination, while a stable number of infections is recorded for chemoprophylaxis + vaccination (Figures 4A and S4A-C, top). The number of infected individuals is consistently lowest for chemoprophylaxis + vaccination. Once, 80% or more of the population is vaccinated there is no difference between the intervention strategies. Overall, a vaccination coverage of over 80% suffices to prevent an outbreak and no additional therapies are required. A similar trend can be observed for the effects of the vaccination coverage on the number of individuals shedding resistant virus (Figures 4A and S4B-C, bottom).

### Effect of vaccine effectiveness on resistance formation is limited

The effectiveness of influenza vaccines varies from season to season due to changes in viral strains. We explored the impacts of different levels of vaccine effectiveness, both with and without the joint use of antiviral mono therapies. Our simulations indicate that higher vaccine effectiveness results in fewer infections, particularly when vaccines are used alone or in conjunction with treatment. However, when vaccination is used with chemoprophylaxis, the infection numbers remain almost constant (Figures 4B and S4D-F, top). The number of infected individuals is consistently lowest for chemoprophylaxis + vaccination. Furthermore, as vaccine effectiveness increases, we observed a decrease in the number of individuals shedding resistant virus (Figures 4B and S4E-F, bottom). Yet, this decrease is again only modest for the joint intervention of chemoprophylaxis and vaccination, and there is a limiting effect when treatment is combined with vaccination. Our analysis indicates that aiming for vaccine effectiveness greater than 50% is more crucial for reducing infection numbers rather than minimizing the emergence of resistance. In summary, our findings highlight that vaccination coverage is the better target to reduce both the total number of infections and of individuals shedding resistant virus.

## Discussion

Public health decisions about the use of antivirals on a large scale should be informed by assessments of the emergence of drug-resistant viral strains and their epidemiological significance. We extended a previously developed mathematical model to capture the effects of drug resistance on the transmission dynamics of influenza viruses and evaluated mitigation strategies such as mono/combination treatment and chemoprophylaxis using antivirals oseltamivir or zanamivir in a non-vaccinated or partially vaccinated population with respect to the potential of drug resistance development.

Our results demonstrate that chemoprophylaxis outperforms treatment in reducing the burden of influenza, both with respect to overall infections and drug resistance formation. Only for high relative transmission rates of the resistant viral strain and when considering resistance formation, did the model predict treatment to outperform chemoprophylaxis. Contrary to treatment, chemoprophylaxis led to a resistance-driven secondary infection wave under high relative transmission rates of the resistant viral strain. Mono- and combination therapy led to similar numbers of infections, however, combination therapy strongly reduced resistance formation. Moreover, our findings demonstrate that a vaccination coverage of at least 80% of the reference population could be sufficient to prevent an infection outbreak altogether, underscoring the significance of vaccination as a primary preventive measure. Overall, our study offers critical insights for the development of effective mitigation strategies for reducing the overall disease burden while considering the risks of drug resistance during influenza outbreaks.

Similar to our findings, earlier models of influenza infection dynamics using M2 protein or NA inhibitors as antiviral monotherapy have shown that chemoprophylaxis leads to an more pronounced and immediate reduction in the number of infections, whereas this effect is more gradual for treatment [9, 10, 11]. Additionally, these models have demonstrated that antiviral monotherapy results in a higher number of individuals shedding resistant virus compared to chemoprophylaxis. Regoes and Bonhoeffer have also noted that high relative resistance transmission rates can lead to an increased emergence of resistance, particularly when chemoprophylaxis is used. Our analysis has now identified that the increased emergence of resistance under these conditions is driven by a secondary wave of infections caused by resistant virus. We provide a quantitative explanation for this phenomenon by deriving and calculating control reproduction numbers. Specifically, our analysis suggests that relative resistance transmission rates above 0.44 can lead to a resistance-driven secondary infection wave in a completely susceptible population. This is consistent with another study, which proposed that relative resistant transmission rates above 0.4 can lead to an uncontrollable pandemic [21]. However, a recent meta-study provides evidence that questions the effectiveness of antivirals for treatment of influenza [22, 23].

We found a vaccination coverage in the reference population of at least 80% to be sufficient to prevent influenza outbreaks. This is in accordance with the critical vaccination coverage when accounting for an imperfect vaccination effectiveness [24]. In 2003 the World Health Organization recommended that EU/EEA Member States aim to achieve an influenza vaccination coverage of at least 75% among high-risk groups [25]. Moreover, we found a vaccination coverage of 80% to result in similar outbreak outcomes than chemoprophylaxis, minus the risk of resistance emergence, as previously reported by Longini et al. [13].

Our results have important implications for the development of effective strategies to prevent and mitigate the impact of influenza infections and the emergence of drug resistance. One of the key challenges in monitoring influenza outbreaks is the high proportion of asymptomatic infections, making it difficult to track the true extent of the outbreak. Our model recapitulates a low fraction of detectable infections, i.e., individuals with symptoms, with at most 0.5 across all investigated scenarios (Figures S1F and S3F). However, the fraction of detectable infections involving resistant virus is considerably higher in our simulations, indicating that it may be possible to monitor resistance emergence reasonably well, if sufficient sequencing efforts would be invested. Next, our analysis identified the relative resistance transmission rate of the resistant viral strain to be critical for the emergence of a resistance-driven secondary infection wave. Overall, infected individuals tend to result in more infections of the same type, e.g., individuals infected with a single-resistant virus tend to lead to more single-resistant infections (Figure S1H). This suggests that monitoring the transmission rates, using methods such as proposed by Leung et al. [26], could help in quantifying the risk of a resistance-driven secondary infection wave early on during an outbreak and enable public health officials to adjust their intervention strategies appropriately. Finally, our results highlight the importance of vaccination in reducing the overall disease burden. While controlling and optimizing vaccine effectiveness is challenging, our model suggests that increasing vaccination coverage is more effective, with a complete prevention of an outbreak for a vaccination coverage above 80% of the reference population. This emphasizes the need for continued investment in vaccine distribution, promoting high vaccination rates among the population.

This study has several limitations. Firstly, our results are only applicable in (semi) closed populations, such as nursing homes, boarding schools, prisons, cruise ships, or hospitals. In an open system, the outcomes of an influenza outbreak could look very differently. Particularly for treatment, relaxing the assumption of a closed population could allow for the emergence of resistance-driven secondary infection waves. Secondly, our model assumes a homogeneously mixed population with similar social behaviors and contact rates across individuals. Behavioral differences among individuals and adaptive behavior, in which susceptible individuals respond to an ongoing outbreak, could considerably influence the model outcomes. To account for behavior, our model would need to be extended to include state- or time-dependent transmissions rates, behaviorally-related compartments, or be reformulated as an agent-based model. Finally, it is important to note that our results primarily focus on the health dimension and do not account for the broader socio-economic costs associated with the evaluated mitigation strategies, including economic costs, potential side effects, and social discomfort. Notably, the number of individuals requiring chemoprophylaxis is nearly three times higher than those requiring treatment, and combination therapy demands twice the amount of antivirals as monotherapy (Figures S1G and S3G). This disparity could be a crucial consideration in crisis management during an influenza outbreak, particularly when the availability of antivirals is limited. Ultimately, whether the increased socio-economic costs for chemoprophylaxis and/or combination therapy are justified will depend on the specific circumstances of each outbreak and require individual assessment.

Future research directions include adapting the model to capture the outcomes of a potential H5N1 avian influenza outbreak, should human-to-human transmission occur. In this scenario, antiviral therapies and zoonotic influenza vaccines will be crucial for prevention and mitigation. A key consideration for effective crisis management will be the anticipation and mitigation of antiviral resistance emergence, especially if considering to administer antiviral chemoprophylaxis broadly across a population and considering initial reports about the detection of potential resistance emergence against oseltamivir among poultry [27].

## Methods

### Model dynamics

#### Influenza infection model without therapy

The influenza model without therapy describes 3 model populations: susceptible individuals 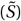, infected but asymptomatic individuals (*Ĩ*), and infected individuals presenting clinical symptoms (*Ĩ*_*s*_) (Figure 1A and C). The dynamics of these model populations are described by the system of ordinary differential equations (ODEs):

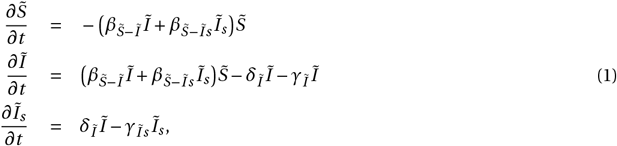

with the initial conditions 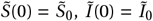, and *Ĩ*_*s*_ (0) = *Ĩ*_*s*0_. Susceptible individuals 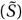 may become infected upon contact with either infected asymptomatic (*Ĩ*) or symptomatic individuals (*Ĩ*_*s*_) with rates 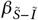 and 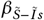, respectively. Once infected, individuals recover at a rate of *γ*_*Ĩ*_ for asymptomatic and *γ*_*Ĩs*_ for symptomatic infections. The rate at which asymptomatic infected individuals develop symptoms is represented by *δ*_*Ĩ*_. Detailed descriptions of the model variables and parameters are provided in Tables 1 and 2, respectively, and the full list of model assumptions is given in the Supplementary Information.

**Table 1:**
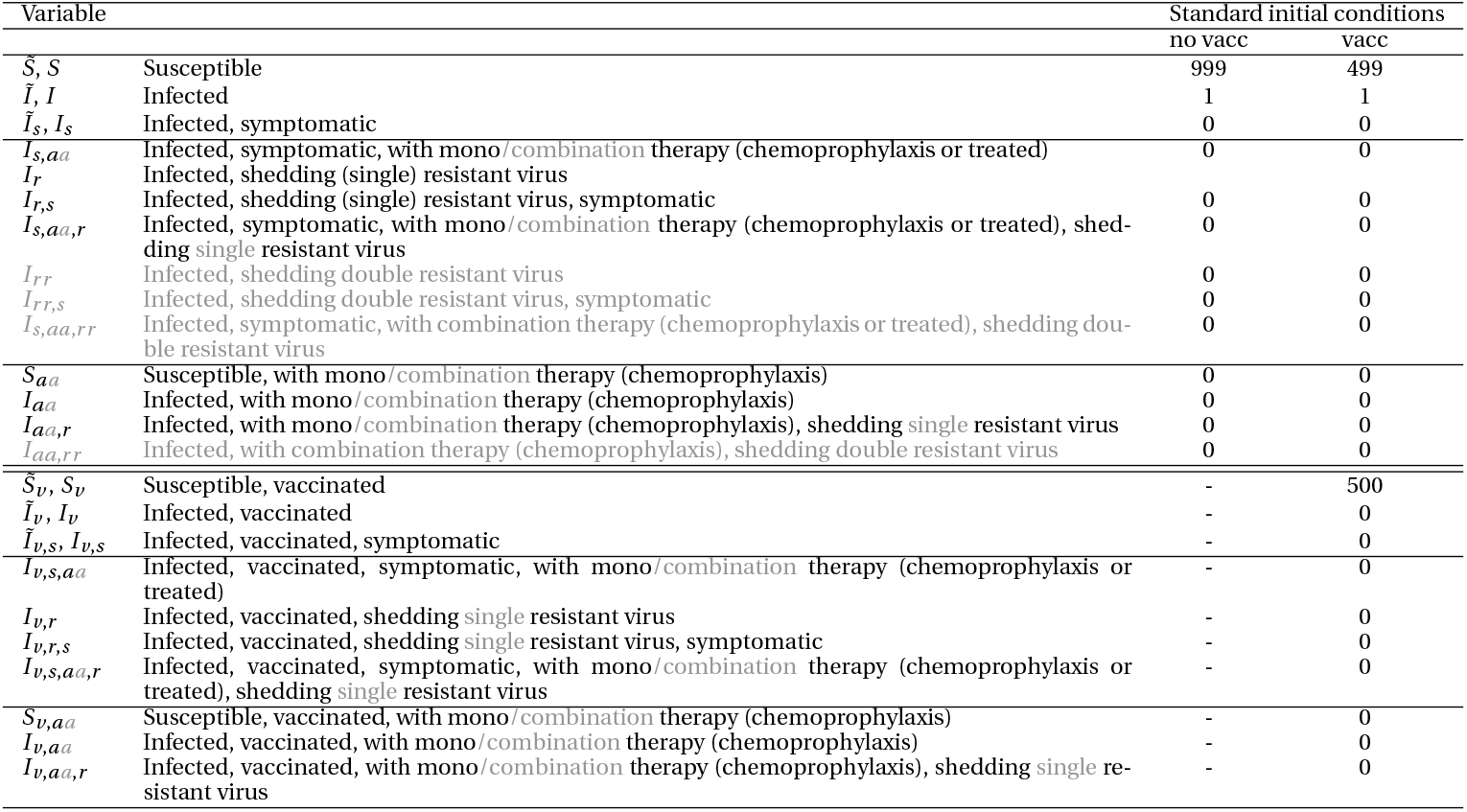
Model variables and initial conditions.

**Table 2:** Parameter values.

| Parameter |  | Standard value (/day) | Reference |
| --- | --- | --- | --- |
| $\gamma_{\tilde{I}} = \gamma_I$ | Basal recovery rate of $I$ | 0.5 | [9] |
| $\gamma_{\tilde{I}_s} = \gamma_{I_s}$ | Basal recovery rate of $I_s$ | 0.25 | [9] |
| $r_{aa} = r_v = r_{v,aa}$ | Relative recovery due to antiviral therapy, vaccination, or both if infected is asymptomatic | 2 | [9] |
| $r_{aa,s} = r_{v,s} = r_{v,aa,s}$ | Relative recovery due to antiviral therapy, vaccination, or both if infected is presenting clinical symptoms | 1.5 | [10] |
| $\delta_{\tilde{I}} = \delta_I$ | Basal rate of symptom development (transition between $I$ and $I_s$) | 0.5 | [9, 28] |
| $d_{aa} = d_v = d_{v,aa}$ | Relative symptom development due to antiviral therapy, vaccination, or both | 0.2 | [9, 28] |
| $\beta_{\tilde{S}-\tilde{I}} = \beta_{S-I}$ | Basal transmission rate between $I$ and $S$ | $2.145 \cdot 10^{-4}$ | [9, 10]* |
| $\beta_{\tilde{S}-\tilde{I}_s} = \beta_{S-I_s}$ | Basal transmission rate between $I_s$ and $S$ | $2.145 \cdot 10^{-3}$ | [9, 10]* |
| $p_{Iaa}$ | Relative transmission if infected was (prophylactically) treated with antiviral mono/combination therapy | 0.67 | [9, 11] |
| $p_{Saa} = p_{Sv} = p_{\tilde{S}v}$ | Relative transmission if susceptible was treated with mono/combination chemoprophylaxis or vaccinated | 0.5 | [10, 28] |
| $p_{Ir}$ | Relative transmission if infected sheds single or double resistant virus | 0.1 | [10, 21] |
| $\theta_{aa}$ | Rate of antiviral mono/combination treatment | 0.7 | [9] |
| $\kappa q$ | Rate of acquired drug resistance if infected is asymptomatic | 0.00823 | ** |
| $\kappa q_s$ | Rate of acquired drug resistance if infected is symptomatic | 0.0823 | ** |

#### Influenza infection model with antiviral mono and combination treatment

The above model was expanded by Stilianakis et al. to describe the population dynamics under antiviral mono treatment [9]. We further expanded this model to also account for antiviral combination treatment (Figure 1A).

The full system of ODEs is given by the following equations:

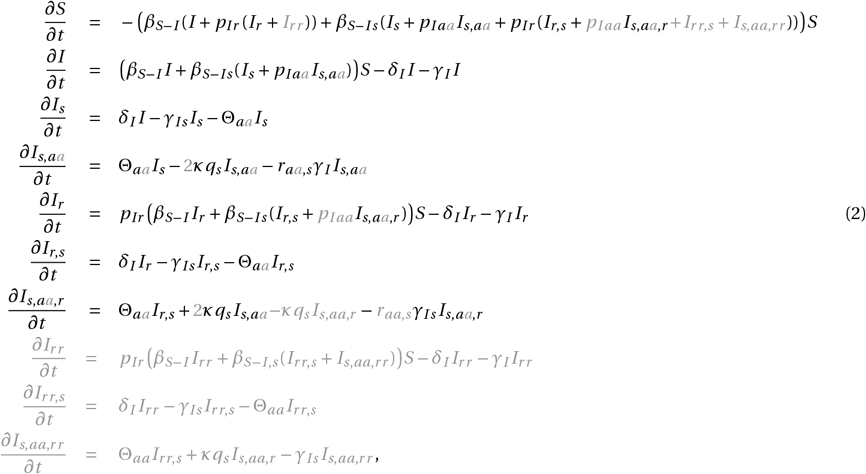

with the initial conditions 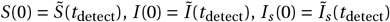 for the treatment-independent states. The time point of outbreak detection and treatment initiation is denoted by *t*_detect_, with *t*_detect_ ≥ 0. The initial conditions for the treatment-dependent states are *I*_*s,aa*_ (0) = *I*_*r*_ (0) = *I*_*r,s*_ (0) = *I*_*s,aa,r*_ (0) = *I*_*r r*_ (0) = *I*_*r r,s*_ (0) = *I*_*s,aa,r r*_ (0) = 0. The new variables and parameters accounting for combination treatment are indicated in gray. Upon detection of an outbreak, symptomatic individuals (*I*_*s*_) receive treatment in the form of either a single (mono) or a combination of two antiviral drugs at a rate Θ_*aa*_, resulting in the state *I*_*s,aa*_. Treatment leads to the emergence of resistant virus at rates 2*κq*_*s*_ and *κq*_*s*_ for single and double resistance under mono and combination treatment, resulting in the state *I*_*s,aa,r*_ and in the case of combination treatment in the additional state *I*_*s,aa,r r*_. We assumed that resistance develops independently against each antiviral drug during combination therapy, leading to a doubling of the resistance formation rate for single resistance formation. However, resistance does not come without cost: to become resistant, the virus acquires mutations, which in turn reduces the viral fitness. Thus, the transmission rates of resistant virus, leading to states *I*_*r*_ and *I*_*r r*_, are reduced by a factor of *p*_*Ir*_. Additionally, treatment results in lower transmission rates by a factor of *p*_*Iaa*_ and increased recovery rates by a factor of *r*_*aa,s*_. The effects reducing transmission (*p*_*Ir*_ and *p*_*Iaa*_) are presumed to operate independently. The benefits of treatment are nullified once the virus becomes resistant.

#### Influenza infection model with antiviral mono and combination chemoprophylaxis

Similarly, the initial influenza infection model was expanded by Stilianakis et al. to describe the population dynamics under antiviral mono chemoprophylaxis [9]. We further expanded this model to also account for antiviral combination chemoprophylaxis (Figure 1C). The full system of ODEs is given by the following equations:

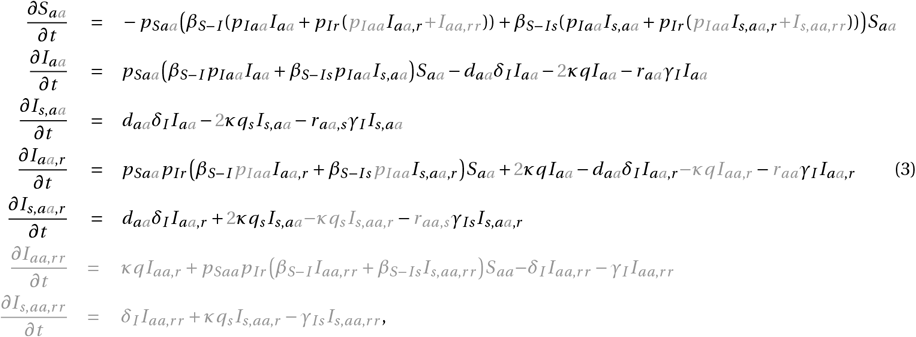

with the initial conditions 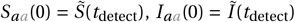, and *I*_*s,aa*_ (0) = *Ĩ*_*s*_ (*t*_detect_) for the states without resistance. The time point of outbreak detection and treatment initiation is denoted by *t*_detect_, with *t*_detect_ ≥ 0. The initial conditions for the resistant states are *I*_*aa,r*_ (0) = *I*_*s,aa,r*_ (0) = *I*_*aa,rr*_ (0) = *I*_*s,aa,r r*_ (0) = 0. The new variables and parameters accounting for combination treatment are indicated in gray. Upon detection of an outbreak, prophylactic treatment is administered to all susceptible (*S*), infected (*I*), and symptomatically infected (*I*_*s*_) individuals. We assume the effects of chemoprophylaxis to be immediate, reducing the transmission rate to susceptible individuals by a factor of *p*_*Saa*_ and the transmission by infected individuals under prophylactic treatment by a factor of *p*_*Iaa*_. chemoprophylaxis leads to the emergence of resistant virus at rates 2*κq*_*s*_ and *κq*_*s*_ for single and double resistance under mono and combination chemoprophylaxis, resulting in the states *I*_*aa,r*_ and *I*_*s,aa,r*_ and in the case of combination chemoprophylaxis in additional states *I*_*aa,rr*_ and *I*_*s,aa,r r*_. Similar to the influenza infection model with antiviral treatment, the transmission rates of resistant virus, leading to states *Ir* and *Ir r*, are reduced by a factor of *p*_*Ir*_, the effects reducing transmission (*p*_*Saa*_, *p*_*Iaa*_, and *p*_*Ir*_) are presumed to operate independently, and the benefits of treatment are nullified once the virus becomes resistant. Additionally, chemoprophylaxis reduces the rate of symptom development by a factor of *d*_*aa*_ and increases the recovery rate of infected asymptomatic and symptomatic individuals by factors *r*_*aa*_ and *r*_*aa,s*_, respectively.

#### Influenza infection model with vaccinated subpopulation without therapy

We further expanded the influenza infection model to describe the effects of vaccination on the population dynamics (Figure 3A and C). The full system of ODEs is given by the following equations:

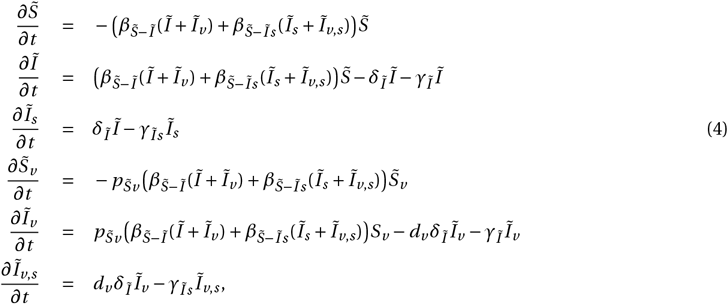

with the initial conditions 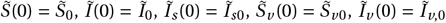, and *Ĩ*_*v,s*_ (0) = *Ĩ*_*v,s*_0. We assumed that a defined fraction of the total susceptible population is vaccinated and that no new vaccinations are administered during the outbreak. The dynamics of the vaccinated subpopulation is similar to that of the non-vaccinated subpopulation, with two distinctions: the rate of virus transmission to vaccinated susceptible individuals is reduced by a factor of 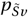 and the rate at which vaccinated individuals develop symptoms is decreased by a factor of *dv*.

#### Influenza infection model with vaccinated subpopulation and antiviral mono treatment

We expanded the influenza infection model with vaccination to also account for the effects of antiviral mono treatment (Figure 3A). The full system of ODEs is given by the following equations:

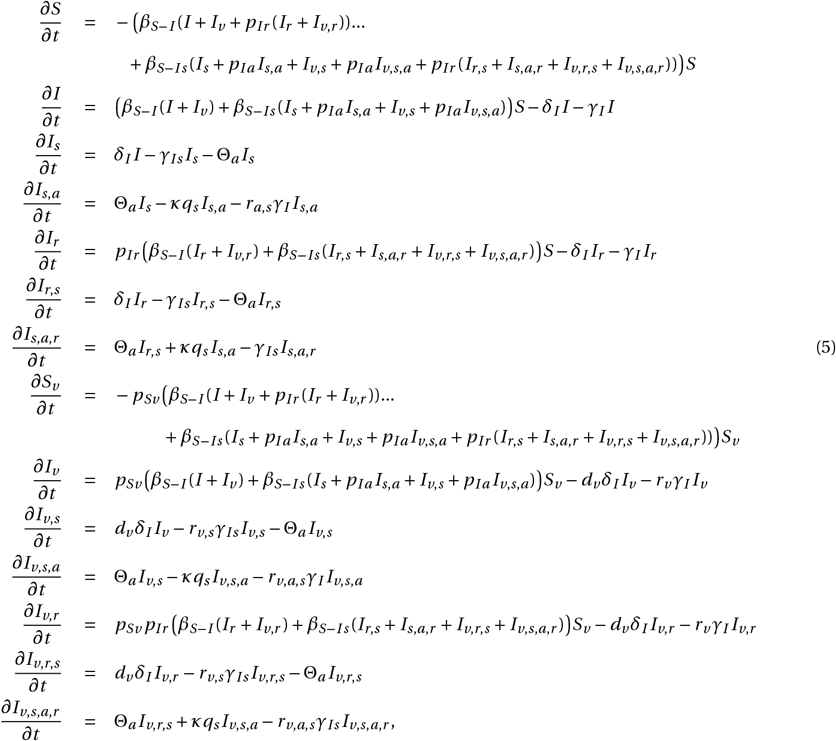

with the initial conditions 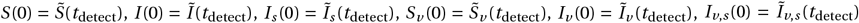 for the treatment-independent states. The time point of outbreak detection and treatment initiation is denoted by *t*_detect_, with *t*_detect_ ≥ 0. The initial values for the treatment-dependent states are *I*_*s,a*_ (0) = *I*_*r*_ (0) = *I*_*r,s*_ (0) = *I*_*s,a,r*_ (0) = *I*_*v,s,a*_ (0) = *I*_*v,r*_ (0) = *I*_*v,r,s*_ (0) = *I*_*v,s,a,r*_ (0) = 0. Again, the dynamics of the vaccinated subpopulation is similar to that of the non-vaccinated subpopulation, with three distinctions: the rate of virus transmission to vaccinated susceptible individuals is reduced by a factor of *p*_*Sv*_, the rate at which vaccinated individuals develop symptoms is decreased by a factor of *d*_*v*_, and the rate at which vaccinated, vaccinated symptomatic, and treated + vaccinated symptomatic individuals recover is increased by the factors *r*_*v, rv,s*_, and *r*_*v,a,s*_, respectively. However, the recovery rate is assumed to be increased jointly and equally by vaccination and treatment, with *r*_*a,s*_ = *r*_*v,s*_ = *r*_*v,a,s*_.

#### Influenza infection model with vaccinated subpopulation and antiviral mono chemoprophylaxis

Similarly, we expanded the influenza infection model with vaccination to also account for the effects of antiviral mono chemoprophylaxis (Figure 3C). The full system of ODEs is given by the following equations:

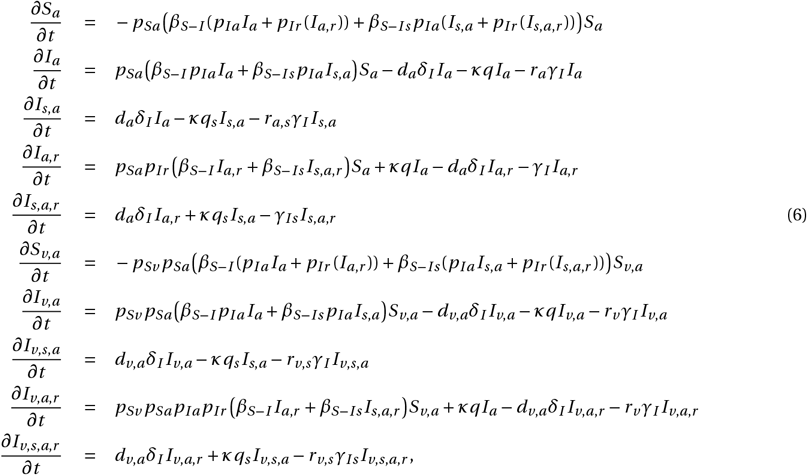

with the initial conditions 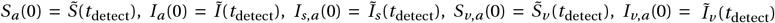 and *I*_*v,s,a*_ (0) = *Ĩ*_*v,s*_ (*t*_detect_) for the states without resistance. The time point of outbreak detection and treatment initiation is denoted by *t*_detect_, with *t*_detect_ ≥ 0. The initial conditions for the resistant states are *I*_*a,r*_ (0) = *I*_*s,a,r*_ (0) = *I*_*v,a,r*_ (0) = *I*_*v,s,a,r*_ (0) = 0. Again, the dynamics of the vaccinated subpopulation is similar to that of the non-vaccinated subpopulation. The rate of symptom development is assumed to be increased jointly and equally by vaccination and chemoprophylaxis, with *d*_*a*_ = *d*_*v*_ = *d*_*v,a*_.

### Model variables

All model variables introduced in the model descriptions and corresponding initial conditions are summarized and explained in Table 1. The additional model variables accounting for combination therapy are highlighted in gray.

### Parameterization

All model parameters introduced in the model descriptions are summarized and explained in Table 2. The additional model parameters accounting for combination therapy are highlighted in gray.

* According to Regoes et al. [10] and Stilianakis et al. [9], we chose the transmission rates such that the basic reproduction number *R*_0_ under the influenza infection model without therapy is:

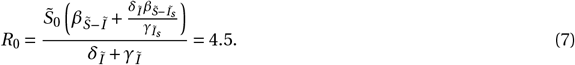

For the resistant strain, this parametrization results in a basic reproduction number of 0.45 under the influenza infection model with antiviral mono treatment.

** According to Kiso et al., 18% of individuals develop de-novo resistance upon treatment with oseltamivir [29]. Similar to Regoes et al. [10], we calculated the rate of acquired de-novo drug resistance under the influenza infection model with antiviral mono treatment, such that:

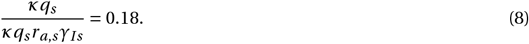

This results in a de novo resistance rate of 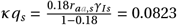. For asymptomatic individuals, we assumed the rate of acquired drug resistance *κq* to be reduced by a factor of 0.1 [9].

### Simulations

#### Simulations with standard values

The simulations are initiated by the infection model without therapy, either with or without a vaccinated subpopulation, initial conditions as given in Table 1, and standard values as given in Table 2. Identification of an outbreak occurs once ∼ 1 − 5% of the population has been infected [9]. For simulations with antiviral therapy, we thus determined the time point, *t*detect, at which 5% of the population were infected and identified as such, i.e. presenting clinical symptoms. Upon detection of an outbreak, treatment is administered to symptomatic individuals at rate Θ_*aa*_, while chemoprophylaxis is administered to all susceptible, infected, and symptomatically infected individuals. We assumed the effects of chemoprophylaxis to be immediate. The simulations are then continued under the respective therapy models up to 40 days since first infection (Figures 1A and C, 3A and C, S2A-D and F-I, and S4). Detailed descriptions of the model variables and parameters are provided in Tables 1 and 2, respectively, and the full list of model assumptions and parameter dependencies is given in the Supplementary Information.

#### Simulations with varying relative de novo resistance formation rates

We simulated the infection dynamics for varying relative de novo resistance formation rates (Figure S2A-D). The simulations were initiated as described in the previous section. However, we adjusted the de novo resistance formation rate *κqs* for the infection model with treatment and *κq* and *κqs* for the infection model with chemoprophylaxis to take on values ranging from 0 (representing no de novo resistance formation) to 10 times their standard values (indicating 69% of individuals undergoing mono treatment develop de novo resistance). The standard value of the relative de novo resistance formation rate is 1. We simulated the outbreak for a total of a 100 days.

#### Simulations with varying relative resistance transmission rates

Similarly, we modeled the infection dynamics for varying relative transmission rates of the resistant virus in comparison to the transmission rate of the wildtype virus (Figure S2F-I). The parameter *p*_*Ir*_, which defines the relative transmission for all resistant populations, was modified to span values from 0 (indicating no transmission of resistance) to 1 (representing a transmission rate equivalent to that of the wildtype virus, implying no fitness loss because of resistance). The standard value of the relative resistance transmission rate is 0.1.

#### Simulations with varying vaccination coverage

We simulated the infection dynamics for varying vaccination coverages, ranging from 0 (representing no vaccinated subpopulation) to 1 (indicating a fully vaccinated population) (Figure S4A-C). We adjusted the initial conditions for the model with vaccination (but without therapy), determined the time of the outbreak detection, and continued the simulations using the mono treatment or chemoprophylaxis model as before. The standard vaccination coverage is 0.5.

#### Simulations with varying vaccine effectiveness

We simulated the infection dynamics for varying vaccine effectiveness, ranging from 0 (representing no vaccine effects) to 1 (indicating maximal vaccine effectiveness) (Figure S4D-F). The overall vaccine effectiveness is an indicator for the joint modification of several model parameters affected by vaccination: *rv, rv,s, rv,a, rv,a,s*, and *dv* (Table 3).

**Table 3:** Model parameters for varying vaccine effectiveness.

| Parameter | Vaccine effectiveness |  |  |  |  |  |  |  |  |  |  |
| --- | --- | --- | --- | --- | --- | --- | --- | --- | --- | --- | --- |
|  | 0 | 0.1 | 0.2 | 0.3 | 0.4 | 0.5 | 0.6 | 0.7 | 0.8 | 0.9 | 1 |
| $r_v$ | 1 | 1.2 | 1.4 | 1.6 | 1.8 | 2 | 2.2 | 2.4 | 2.6 | 2.8 | 3 |
| $r_{v,s}$ | 1 | 1.1 | 1.2 | 1.3 | 1.4 | 1.5 | 1.6 | 1.7 | 1.8 | 1.9 | 2 |
| $r_{v,a}$ | 2 | 2 | 2 | 2 | 2 | 2 | 2.2 | 2.4 | 2.6 | 2.8 | 3 |
| $r_{v,a,s}$ | 1.5 | 1.5 | 1.5 | 1.5 | 1.5 | 1.5 | 1.6 | 1.7 | 1.8 | 1.9 | 2 |
| $d_v$ | 1 | 0.72 | 0.53 | 0.38 | 0.28 | 0.2 | 0.15 | 0.11 | 0.08 | 0.06 | 0.04 |

The lower limits of the vaccine-dependent parameters are chosen such that they equal the parameters without vaccination (e.g. *r*_*v*_ = 1 and *r*_*v,a*_ = *r*_*a*_). The upper limits of the vaccine-dependent model parameters are chosen freely. The standard vaccine effectiveness is 0.5. The recovery is assumed to increase jointly and equally by vaccination and treatment. Thus, we adapted *r*_*v,a*_ and *r*_*v,a,s*_ only when the reduction of the recovery rate by therapy, *r*_*a*_, and *r*_*a,s*_, respectively, is exceeded. By definition, this occurs only when the vaccine effectiveness is larger than 0.5.

### Outcome of outbreak

To summarize the outcome of an outbreak, we determined the:

- number of remaining susceptible individuals after the outbreak (Figures 1E and 3E)
- total number of individuals who got infected during the outbreak (Figures 1E and 3E)
- total number of individuals who presented clinical symptoms during the outbreak (Figures S1E and S3E)
- total number of individuals who shed resistant virus during the outbreak (Figures 1E and 3E)
- total number of individuals who presented clinical symptoms and shed resistant virus during the outbreak (Figures S1E and S3E)
- total number of individuals who received therapy (Figures S1G and S3G).

The model populations contributing to each of these summary populations are shown in Table S2 for each model. The number of remaining susceptible individuals after the outbreak is given by *S*(*t*_end_) and *S*(*t*_end_) + *S*_*v*_ (*t*_end_) for models without and with vaccination, respectively, and *S*_*aa*_ (*t*_end_) or *S*_*aa*_ (*t*_end_)+ *S*_*v,aa*_ (*t*_end_) for models with chemoprophylaxis and vaccination + chemoprophylaxis, respectively, where *t*end = 40 is the considered duration of the outbreak. For the other summary populations, we summed up the integrals of all inflows. More specifically, we first determined the time-resolved flow of each model process. Then, using the MATLAB function *trapz*(*t*,.), where *t* is a vector of the simulated time points and ‘.’ is the previously determined flow for a specific process, we approximated the respective integral (Figures S1A and C and S3A and C) [30]. Finally, we summed over all the integrals representing an inflow into the summary population. For example, the number of infected individuals in the model with mono treatment is determined by two processes: 1) the infection of susceptible individuals with wildtype virus and 2) the infection of susceptible individuals with resistant virus. The time-resolved flows of the two infection processes are given by:

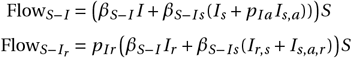

and the overall number of infected individuals *ℐ* is given by:

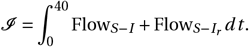

The integral is approximated with *trapz*(*t*, Flow*S*−*I*) = 895 and 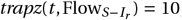, with *t* = 0 : *t*end. Thus, the total number of individuals who got infected during the outbreak with mono treatment is given by 895 + 10 + 1 = 906, including the initial infected individual. Processes involving treatment, are only considered for time points after treatment initiation, *t*_detect_. The number of individuals treated with chemoprophylaxis is given by *S*_*aa*_ (0) and *S*_*aa*_ (0) + *S*_*v,aa*_ (0) for models without and with vaccination. Using the summary populations, we determined the fraction of detectable infections and resistant detectable infections (Figures S1F and S3F).

### The control reproduction number

The basic reproduction number, represented as *R*_0_, quantifies the average number of secondary infections produced by a single infected individual in a completely susceptible population, serving as an index of a disease’s transmission capability. When *R*_0_ exceeds 1, the disease is expected to propagate, whereas an *R*_0_ below 1 indicates that the disease will be extinguished over time. Conventionally, the calculation of *R*_0_ focuses on the number of new cases of population *x* caused by an infectious individual of population *x*. This measure does not provide information about the number of new infections of type *y* resulting from an infectious individual belonging to a different population *x*. To address this gap and better assess the impact of treatment and the emergence of resistance, we introduced the control reproduction number, *R*_*x*−*y,z*_ [31]. This metric gives the number of individuals of population *y*, that are produced when placing an individual of population *x* into a fully susceptible population under the model dynamics *z*, where *z* ∈ {none, treat, proph}. Specifically, the values 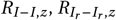, and 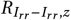, within the context of model dynamic *z*, represent the analogous control reproduction numbers for drug-susceptible, single-resistant, and double-resistant strains, respectively. In the scenario where no treatment is administered, the basic reproduction number is calculated as follows:

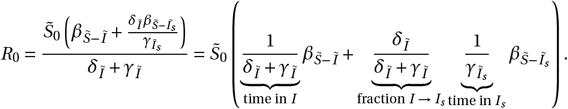

The basic reproduction number can be understood as an aggregated value, calculated by summing the product of three components for each infectious compartment: the proportion of individuals that transition into the compartment (influenced by the rates of inflow and outflow from preceding compartments), the duration of infectiousness within the compartment (inversely related to the outflow rate of that compartment), and the specific transmission rates associated with the compartment. Similarly, we derived the corresponding control reproduction numbers by considering the aforementioned elements (transitional proportions, durations of stay in infectious compartments, and transmission rates) under the model dynamics *z* for *x* = *I* or *x* = *I*_*aa*_ :

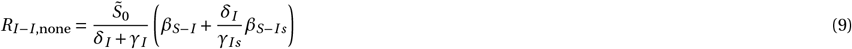

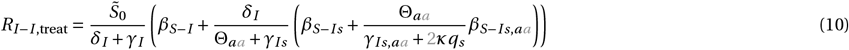

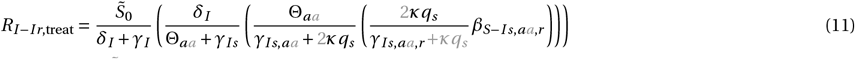

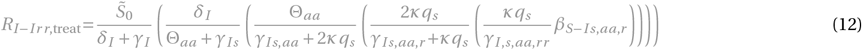

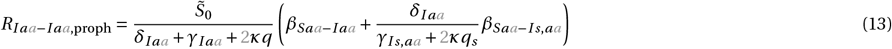

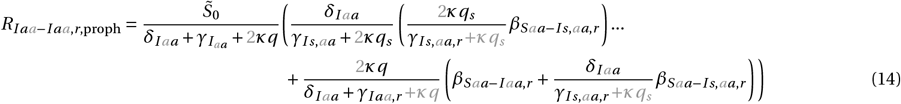

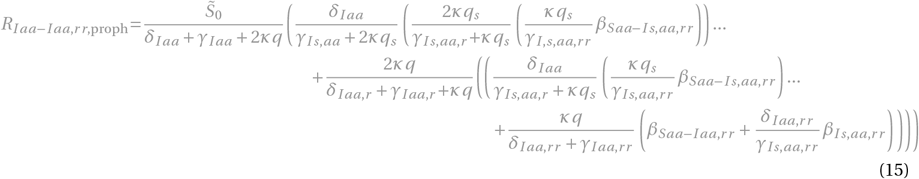

for *x* = *I*_*r*_ or *x* = *I*_*aa,r*_ :

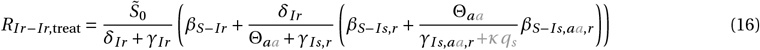

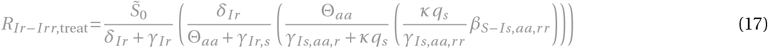

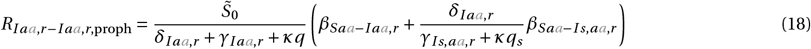

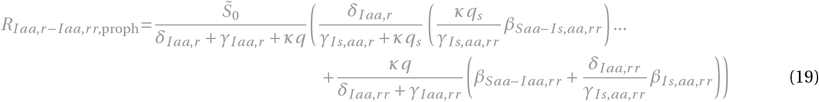

for *x* = *I*_*r r*_ or *x* = *I*_*aa,rr*_ :

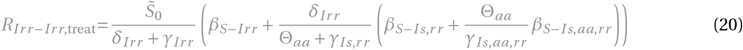

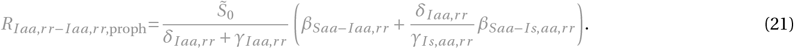

The terms and control reproduction numbers accounting for combination therapy are marked in gray. The control reproduction numbers for the standard model parameterization are summarized in Figure S1H and Table S3.

#### Control reproduction numbers for varying relative de novo resistance formation rates

We computed the control reproduction numbers for varying relative de novo resistance formation rates (Figure S2E). Rates *κq* and *κq*_*s*_ were adjusted to take on values ranging from 0 to 10 times their standard values in equations (11) and (12) for mono and combination treatment, respectively, and equations (14) and (15) for mono and combination chemoprophylaxis, respectively. The standard value of the relative de novo resistance formation rate is 1 (Table 2).

#### Control reproduction numbers for varying relative resistance transmission rates

We computed the control reproduction numbers for varying relative resistance transmission rates (Figure S2J). The relative resistance transmission factor *p*_*Ir*_ was adjusted to span values from 0 to 1 in equations (11) and for mono and combination treatment, respectively, and equations (14) and (15) for mono and combination chemoprophylaxis, respectively. The standard value of the relative resistance transmission factor is 0.1 (Table 2). The factor *p*_*Ir*_ determines the transmission rates for resistant model populations (e.g. *β*_*S*−*Irr*_, *β*_*S*−*Irr, s*_, *β*_*S*−*Iaa,rr*_, *β*_*S*−*Is,aa,rr*_, *β*_*Saa*−*Iaa,rr*_, and *β*_*Saa*−*Is,aa,rr*_). More details on the relations between parameters is summarized in Table S1.

#### Control reproduction number for *p*_*Ir*_ = 1

For a fixed relative resistance transmission factor *p*_*Ir*_ = 1 (indicating no fitness loss and a transmission rate equal to that of the wildtype virus), we computed the control reproduction number. The control reproduction number here relaxes the assumption of having a fully susceptible population 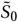 and instead accounts for the decreasing number of susceptible individuals throughout an outbreak (Figure S2K). Using the simulations, we determined the time-dependent model variables *S* and *S*_*aa*_ under the mono and combination treatment model dynamics and substituted 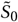 in equations (11) and (12) for mono and combination treatment, respectively, and in equations (14) and (15) for mono and combination treatment, respectively. We want to note, that before the outbreak detection and therapy initiation/administration at *t*_detect_, no resistance is formed and the control reproduction number is equal to zero.

### Implementation

The analysis was performed with MATLAB 2023a.

## Data Availability

There is no original data underlying this work.

## Author contributions

Conceptualization, L.S., N.I.S.; Data curation, L.S.; Formal analysis, L.S.; Funding acquisition, N/A; Investigation, L.S.; Methodology, L.S., N.I.S.; Project administration, N.I.S.; Resources, N.I.S.; Software, L.S.; Supervision, N.I.S.; Validation, L.S.; Visualization, L.S.; Writing – original draft, L.S., N.I.S.; Writing – review and editing, L.S., N.I.S.

## Acknowledgments

The views expressed are purely those of the authors and may not in any circumstances be regarded as stating an official position of the European Commission. The authors used GPT@JRC to improve the language and writing style during the drafting of this manuscript. Afterwards, the authors reviewed and edited the sections as needed and take full responsibility for its content.

## Data and code accessibility

There is no original data underlying this work. The MATLAB code corresponding to this manuscript will be made available via GitHub upon publication.

